# Large-scale blood mitochondrial genome-wide study provides novel insights into mitochondrial disease-related traits

**DOI:** 10.1101/2023.06.12.23291273

**Authors:** S Cannon, T Hall, G Hawkes, K Colclough, RM Boggan, CF Wright, SJ Pickett, AT Hattersley, MN Weedon, KA Patel

## Abstract

**Background/Objectives:** Whole genome sequencing (WGS) from large cohorts enables the study of mitochondrial DNA (mtDNA) variation on human health. We aimed to investigate the influence of common, rare, and pathogenic mtDNA variants on 15 mitochondrial disease-related phenotypes.

**Methods:** Using WGS from 179,862 individuals from in the UK Biobank, we identified mtDNA variants using MitoHPC. We performed extensive association analyses with 15 mitochondrial disease-relevant phenotypes. We compared the results for the m.3243A>G variant with those from a clinically referred patient cohort.

**Results:** Of 15,881 mtDNA variants, 12 homoplasmic and one heteroplasmic variant had genome-wide significant associations. All homoplasmic variants increased aspartate aminotransferase level and three were novel, low frequency, variants (MAF∼0.002 and beta∼0.3 SD). Only m.3243A>G (MAF=0.0002) associated with diabetes (OR=5.6, 95%CI [3.2-9.9]), deafness (OR=12.3, 95%CI [6.2-24.4]) and heart failure (OR=39.5, 95%CI [9.76-160.1]). Multi-system disease risk and penetrance of all three traits increased with m.3243A>G level. Diabetes risk was further influenced by common nuclear genome variation. The penetrance of diabetes with m.3243A>G in the UK Biobank was lower than clinically referred patients, partly attributed to lower heteroplasmy. Of 73 pathogenic mitochondrial disease variants, most were rare in the population with low penetrance.

**Conclusion:** Our study highlights the utility of WGS for investigating mitochondrial genetics within a large, unselected population. We identified novel associations and demonstrated that pathogenic mitochondrial variants have lower penetrance in clinically unselected than clinically referred settings. m.3243A>G associated with mitochondrial-related phenotypes at higher heteroplasmy. Our findings suggest potential benefits of reporting incidentally identified m.3243A>G at high heteroplasmy levels.

## Introduction

Mitochondrial function is fundamental to human life but can be impaired by pathogenic mitochondrial (mt)DNA variants, leading to disease with variable expressivity and penetrance (1, 2, 3). Studies of predominantly clinically ascertained cohorts have identified >90 pathogenic variants in the mitochondrial genome (1, 2, 4, 5). These variants cause rare heterogeneous mitochondrial disorders, including complex multi-organ disease (1). Multiple studies have also reported mtDNA variants associated with complex diseases such as maternally-inherited diabetes (6), metabolic diseases (7, 8), Parkinson disease (9), neuroticism (10), as well as stroke and psoriasis (11), among others. These variants provide insight into the function of human mitochondria and the pathogenesis underlying these diseases (12).

Previous mitochondrial DNA studies have had limitations. For example, when identifying mitochondrial variants which affect complex traits, studies have either assessed a subset of common variants (n∼700) in large, unselected cohorts (13, 14), or all variants in small, often clinically selected cohorts (n=100-2800) focussing on a certain phenotype (15, 16). Studies of pathogenic variants have been primarily restricted to patients and family members with certain phenotypes, likely resulting in over-inflated penetrance and expressivity estimates as seen for nuclear pathogenic variants (17, 18, 19). Large-scale rare and common mitochondrial genome-wide association studies in clinically unselected populations can overcome these limitations, and lead to novel insights into the impact of mitochondrial variants on human health (20).

The recent availability of whole genome sequencing data (WGS) from large cohorts provides a unique opportunity to study rare and common mitochondrial variants. WGS data have recently been made available in large population cohorts such as the UK Biobank (UKB, n=200,030) (21) and All of Us (22). WGS captures information from both mitochondrial DNA (mtDNA) and nuclear DNA, providing high-quality genotyping for all mitochondrial variations, both rare and common. Importantly, it also provides accurate data on variant heteroplasmy (proportion of mtDNA molecules with alternate allele) and mtDNA copy number (mtCN), making it an ideal technology to study all, and particularly rare, heteroplasmic and homoplasmic mitochondrial variation (23). These large cohort studies also provide excellent opportunity to assess penetrance and expressivity of known pathogenic variants in a clinically unselected population which is crucial before considering the incidental reporting of mitochondrial variants.

Here we use WGS data from a large multi-ethnic population cohort of 200,030 individuals from the UKB. We aimed to identify novel rare and common variants associated with 15 common mitochondrial disease-related traits and assess the prevalence and penetrance of known pathogenic variants in an unselected population cohort.

## Methods

### Study subjects

#### UK biobank

The UKB is an ethically-approved population cohort of ∼500,000 individuals from the UK (24). The UKB contains deep phenotype data from self-reporting, hospital and GP records, and measurements of 30 blood biomarkers including HbA1c and liver enzymes, which are paired with detailed genetic data. Whole exome sequencing and genotyping array imputation data are available in the entire cohort, and at the time of writing (May 2023). WGS data is available on 200,030 participants of diverse genetic ancestries. The lack of selection on any specific disease phenotypes, and large sample size, makes it an ideal cohort to study rare genotype-phenotype associations. Cohort characteristics of individuals included in the current study are summarised in Table S1.

#### Clinically identified probands with m.3243A>G pathogenic variant

We included 95 probands who were referred from routine diabetes clinics in the UK to Exeter Genomics laboratory, Royal Devon University Healthcare Hospital with suspected mitochondrial-related diabetes and found to harbour the m.3243A>G variant. The study was approved by the Wales Research ethics Committee 5 (22/WA/0268). Cohort characteristics for these individuals are summarised in Table S2.

#### Clinical phenotypes

We analysed 15 diseases/traits that have been commonly associated with mitochondrial disease (1) (25) and were possible to generate from the data available in the UKB. We used self-report data, ICD9/10 codes, medication, and biomarkers to find these phenotypes (Table S3).

### Genetic data

#### Whole genome sequencing data

UKB generated WGS alignment files (CRAM format) using the deCODE pipeline as previously described (21). Briefly, 500ng of genomic DNA per-sample underwent paired-end sequencing of 150 base pairs on Illumina NovaSeq6000 sequencers with the S4 flow cell and real time base calling. Data were aligned to GRCh38 before undergoing contamination and data quality control. A detailed description is available from webpage https://biobank.ndph.ox.ac.uk/showcase/label.cgi?id=180. We derived genetic ancestry for Europeans (EUR-like), Africans (AFR-like) and South Asians (SAS-like) via comparison of genotypes derived from the UK Biobank Axiom Array to nuclear genome principal components derived from the 1000 genomes project.

#### Mitochondrial variant calling from whole genome sequencing data

We used MitoHPC with Mutect2 in mitochondrial mode to acquire mitochondrial variants from the WGS CRAM files of the 200,030 participants (26) (27). MitoHPC is specifically designed to detect mitochondrial single nucleotide variants (SNV) in large WGS datasets and provides accurate heteroplasmy estimates by using a consensus mitochondrial sequence for each sample. MitoHPC also provides mitochondrial copy number estimation (as a proportion of reads mapped to the mitochondrial and nuclear genomes), haplogroup determination, sequencing coverage statistics, as well as quality metrics at both sample and variant levels.

We used stringent sample exclusion criteria to minimise false-positive low heteroplasmic variant calls. We excluded potentially contaminated samples (e.g. multiple dominant haplogoups in a single sample n=736), samples with low coverage (min <200x or mean <500x, n=1126), or samples with multiple nuclear DNA of mitochondrial origin (nuMT) variants flagged by MitoHPC (n=17,485) (26). Extremes of mitochondrial copy number (Q1-1.5xIQR and Q3+1.5xIQR, n=13), were also excluded. An additional 821 samples were excluded where low quality, or missing, genotype data excluded them from the generation of a genetic relatedness matrix (n=731) or were not able to be processed by MitoHPC (n=90). In total, Of the 199,209 samples that were processed by MitoHPC, 179,862 (90.3%) samples passed our robust sample filtering.

Our variant level filtering was in line with previous large studies (20). Specifically, we removed low quality variants identified by GATK FilterMutectCalls as well as multiallelic indels and those identified within six known low complexity regions (20) or at 382 nuMT sites determined by MitoHPC (26). This provided us with 15,881 variants in 179,862 individuals. Of these, 12,009 (∼76%) variants had a minor allele frequency <0.01% which would not have been reliably captured using genotyping array technology (28) and 8,896 variants (56%) had minor allele count (MAC) ≥5. Given our stringent filtering criteria, our primary analysis used variants with ≥3% heteroplasmy. We also performed sensitivity analysis for variants with ≥5% and ≥10% heteroplasmy, as shown in Table 1, to remove the possible false association by low level mitochondrial heteroplasmic calls.

**Table 1:** Heteroplasmy (%) thresholds used to define heteroplasmic and homoplasmic variants.

| Definition | Reference genotype | Heteroplasmic | Homoplasmic |
| --- | --- | --- | --- |
| Primary | <3 | $\geq 3$ and <97 | $\geq 97$ |
| Sensitivity | <5 | $\geq 5$ and <95 | $\geq 95$ |
| Sensitivity | <10 | $\geq 10$ and <90 | $\geq 90$ |

#### Type 2 Diabetes (T2D) genetic risk score

We generated a T2D genetic risk score for individuals in the UKB based on 88 T2D associated variants from genotyping array as identified in previous genome wide association studies (29) (30) that did not include the UKB. We also generated the partitioned T2D scores as described by Udler *et al.* (2018)(31).

#### m.3243A>G testing for clinically referred probands

We used digital droplet PCR (ddPCR) on a Bio-Rad (California, USA) machine to analyse blood DNA for presence of and to determine heteroplasmy of m.3243A>G. PCR primer sequences for the m.3243A>G ddPCR assay are described by Singh *et al* (2006)(32). Plates were run on an AutoDG Droplet Generator (Bio-Rad) with DG32 cartridges according to manufacturer’s instructions and analysed using QuantaSoft version 1.7 software (Bio-Rad). All samples were tested in triplicate and droplet data combined for final analysis of heteroplasmy. Heteroplasmy >2% was considered positive.

#### List of Known pathogenic mitochondrial variants

We reviewed all proposed pathogenic variants from the MitoMAP repository (accessed Oct 2022) https://mitomap.org/foswiki/bin/view/MITOMAP/ConfirmedMutations. For this study, we included a variant (n=73, Table S4) if it is curated as pathogenic or likely pathogenic based on the multiple MitoMAP criteria (status = Cfrm) (https://www.mitomap.org/foswiki/bin/view/MITOMAP/MutationsCodingControlCfrm) and where available, annotated as pathogenic or likely pathogenic based on the independent criteria defined by the international ClinGen consortium e.g. https://mitomap.org/cgi-bin/search_allele?variant=7445A>G.

### Statistical analysis

#### Mitochondrial genome wide association

We performed single variant association analysis for variants with a MAC ≥5 (n=8,896) using REGENIE (v3.1.4) (33) for 15 selected disease/traits (Table S3). REGENIE performs genome wide association testing for large sample sizes and is robust to unbalanced case-control ratios, and controls for population structure by utilising a genetic relatedness matrix generated from common, independent nuclear single nucleotide polymorphisms. To generate the genetic relatedness matrix, we used participants who had both a whole genome sequence and whole exome sequence data available.

For variants which were only ever heteroplasmic, or only ever homoplasmic in our cohort, we compared them against the reference genotypes. For variants where some individuals were defined as heteroplamic and some homoplasmic, our primary comparison was individuals with homoplasmic variant versus individuals with heteroplasmic or reference genotype. We performed secondary analysis by comparing heteroplasmic or homoplasmic variant genotypes together versus reference genotypes. We rank inverse normalised the continuous traits and the analysis was adjusted for age at recruitment, sex, sequencing batch, recruitment centre, mitochondrial copy number (mtCN) and the first 40 nuclear genetic principal components (PC). Our significance threshold was the Bonferroni corrected p value of <3.75x10^-7^ calculated by number of variants and traits that were analysed (=0.05/8896/15).

#### Rare variant aggregation testing

We performed aggregate testing of rare mtDNA variants (MAF<0.1%) annotated as either missense, synonymous, or loss-of-function by Ensembl VEP (34) which were grouped by mitochondrial transcript, the d-loop, and 100bp sliding windows at 10bp intervals. Previously known pathogenic variants were also grouped by their known disease phenotypes (Table S4). We used REGENIE to perform burden, SKAT and ACAT aggregate tests (33) for association with all 15 traits. We used a Bonferroni corrected p value of <2x10^-6^ as our significance threshold, calculated by number of masks and traits that were analysed (=0.05/1696/15).

#### Penetrance of diabetes for m.3243A>G

We used Kaplan-Meier survival estimates to estimate the age-dependent penetrance of diabetes. A log-rank test for equality was used to compare the penetrance of diabetes between the groups. Cox’s regression was used to compute the hazard ratio for developing diabetes with or without the adjustment of covariates. The analysis included all individuals without diabetes and individuals with diabetes without missing age at diagnosis (92% of all diabetes cases). We used STATA 16 for all analysis.

## Results

### Mitochondrial-wide single variant association identified associations with diabetes, deafness, heart failure and aspartate aminotransferase

We performed single variant association analyses for 8,896 mitochondrial variants (MAC ≥ 5) with 15 disease/traits that are commonly reported in individuals with mitochondrial disease (Tables S3, S5). In a European-like ancestry analysis, we identified four disorder/traits where variants were associated at our Bonferroni-corrected significance threshold (p<3.75x10**^-7^**) of which enzyme aspartate aminotransferase (AST) was the only trait with >1 variant association. This included 12 homoplasmic variants associated with AST and only one heteroplasmic variant (m.3243A>G) associated with diabetes (MAF=0.00024, OR=5.6, 95% CI [3.2-9.9], P=4x10^-9^) and hearing aid use (OR=12.3, 95% CI [6.2-24.4], P=6x10^-13^ Figure 1A, Table S6). Sensitivity analysis using a higher heteroplasmy threshold (≥10%) observed one additional association of heart failure with the same m.3243A>G variant (OR=39.5, 95% CI [9.76-160.1]; P<3x10^-7^; Table S5). Sensitivity analysis using the dominant model of association (reference vs either heteroplasmic or homoplasmic) did not identify additional variants and showed consistent results.

### Lead AST associated variant associates with other liver enzymes but not non-alcoholic fatty liver disease

Of the 12 primarily homoplasmic variants associated with increased AST levels, three were low frequency novel variants with a large effect size (MAF ∼0.002, β∼0.33) compared to previously known variants (Figure 1, Tables S5, S6, S7). The conditional analysis of each variant for all others showed that the AST associations were led by one previously reported variant: m.15758A>G (MAF=0.02; β=0.09, P=5x10^-8^) (13) and one novel variant: m.13488C>T (MAF=0.002; β=0.33, P=7x10^-11^) (Table S6). m.15758A>G causes a missense change in the MY-CYB gene and m.13488C>T is a synonymous change in the MT-ND5, both components of respiratory chain complexes. We next tested the association of these two lead variants with four other liver biomarkers; alanine aminotransferase (ALT), alkaline phosphatase (ALP), gamma-glutamyl transferase (GGT), bilirubin and, non-alcoholic fatty liver disease (NAFLD). Both these variants associated with increased ALT (beta 0.06 and 0.24 respectively, p<6x10^-4^) but neither were associated with any other liver markers or non-alcoholic fatty liver disease (Table S8).

**Figure 1.**
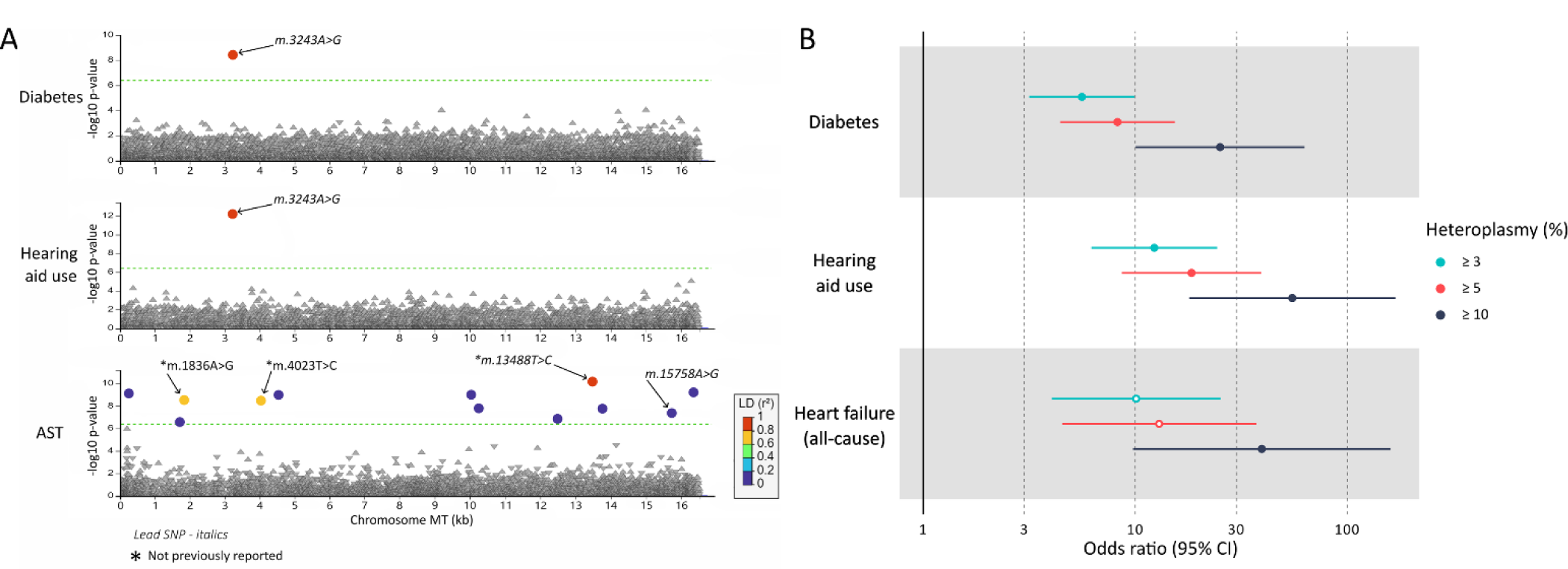
Summary of hypothesis free mitochondrial genome wide associations for 15 mitochondrial disease phenotypes and *m.3243A>G* associations at increasing heteroplasmy thresholds. **A** LocusZoom plot highlighting all significant associations between mitochondrial variants in the UK Biobank with measured heteroplasmy ≥ 3% (n=8,896, MAC≥ 5) and the 15 tested phenotypes. Green dashed line denotes the Bonferroni-corrected significance threshold (P<4.03x10^-7^). R^2^ Correlation is relative to the most significant variant in each plot. **B** Forest plot of *m.3243A>G* associations at increasing minimum heteroplasmy. Solid points denote associations passing the Bonferroni significance threshold (P<3.75x10^-7^). Heart failure (all-cause) was the only phenotype to have a significant association in the sensitivity tests for any traits. AST=aspartate transferase.

### Rare variant aggregation and other ancestry analyses did not identify additional novel associations

We next performed rare variant (MAF <0.001) aggregation burden analyses to determine whether rare mtDNA variants were associated with these mitochondrial-related traits. We aggregated rare variants by mitochondrial transcript and for 100bp sliding windows at 10bp intervals. We did not identify any new associations at a Bonferroni-corrected P value (<2x10^-6^; =0.05/1696/15). We did not identify single or rare variant aggregated associations in an analysis of African-like or South Asian-like ancestry individuals at Bonferroni corrected significance threshold.

### The m.3243A>G association with diabetes, deafness, and heart failure greatly increases with blood heteroplasmy ≥10%

The variant m.3243A>G was associated with diabetes and deafness at ≥3% heteroplasmy and these associations were stronger with higher heteroplasmy. The median m.3243A>G heteroplasmy was 7.8% (IQR 5.3-13.1%) for 83 m.3243A>G carriers (all ancestry) in the UKB. Of which, 23% had 3-4.9%, 47% had 5-9.9% and 30% had ≥10% (Figure S1). Consistent with cross-sectional analysis, we observed higher age-related penetrance of diabetes using Cox proportional hazard model for diabetes (HR 5.8, 95% CI: [3.8-9], p=3.5x10^-15^) in all m.3243A>G carriers compared to non-carriers (Figure S2). Although, the overall risk was higher, this was predominantly driven by individuals with heteroplasmy ≥5% (Figure 2A, Table S9). The risk of diabetes was similar to the background/non m.3243A>G carriers for individuals with m.3243A>G heteroplasmy of 3-4.9% (HR 0.98, 95% CI [0.14-6.9], p>0.9) but increased to 3.8 (95% CI [1.8-8], P=3.9x10^-4^) for 5-9.9%, and further increased to 20.6 (95% CI [11.7-36.4], P=1.2x10^-25^) for individuals with ≥10% (Table S9) (P heterogeneity=0.0001). Of the 16,386 individuals with diabetes, m.3243A>G was responsible for 0.14% of diabetes. All results were consistent for unrelated European-like ancestry individuals and when using tertiles of age-adjusted m.3243A>G heteroplasmy (based on a previously published formula (35)) (Figure S3A, Table S10). These data suggest that m.3243A>G level in the blood is an important factor underlying penetrance in a clinically unselected population.

**Figure 2.**
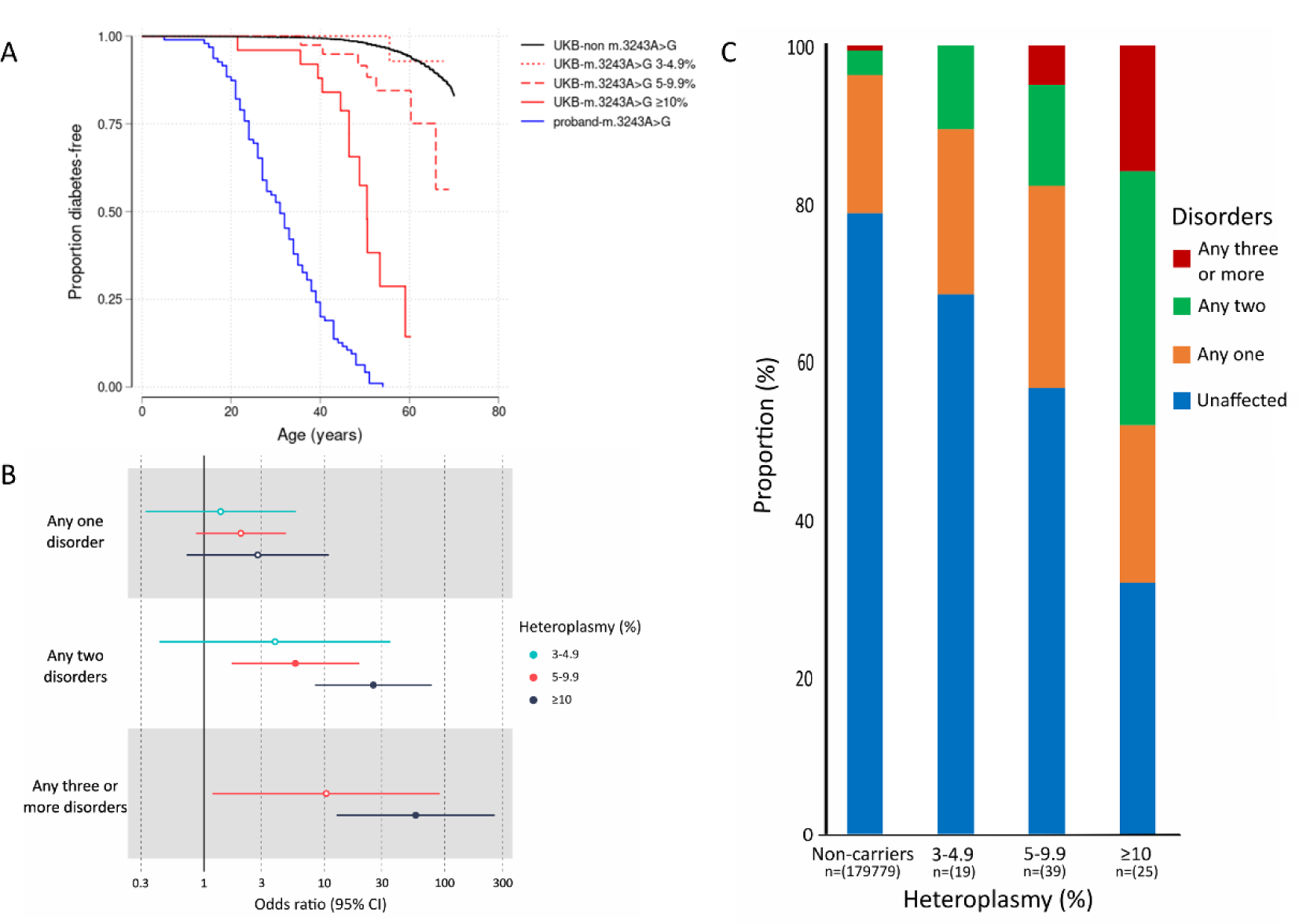
Penetrance of diabetes in *m.3243A>G* carriers in clinical and unselected cohorts and associations with multisystem phenotypic presentations. **A** Kaplan-Meier survival curves of diabetes for m.3243A>G individuals in the UK Biobank (n=83) split by 3-4.9% (n=19), 5-9.9% (n=39) and ≥10% heteroplasmy (n=25) and clinically identified probands with m.3243A>G (n=95). The log rank test p value for these three groups against the noncarriers in the UK Biobank were 0.8, 5x10^-5^ and 9x10^-71^, respectively. The log rank test p value of UKB ≥10% group against proband was 1x10^-9^ **B** Significant multisystem disorders with one or more presenting phenotypes of: heart failure (all-cause) or non-ischaemic cardiomyopathy, constipation, hearing problems, diabetes, stroke, renal disease, and epilepsy. Hollow points denote non-significant associations when adjusting for multiple testing (p<0.0056; p=0.05/9) **C** Proportion of *m.3243A>G* carriers (n=83) with multiple affected systems at increasing heteroplasmy thresholds split by 3-4.9% (N=19), 5-9.9% (n=39) and ≥10% heteroplasmy (n=25).

### m.3243A>G is associated with higher risk of multisystem disorder which increases with higher heteroplasmy

The variant m.3243A>G is commonly associated with a multi-system disorder when ascertained clinically. It is not clear if this occurs when assessed in a hypothesis-free manner in clinically unselected cases, and whether any risk is affected by m.3243A>G levels. To assess this, we grouped mitochondrial-related disorders where we observed the nominal association with m.3243A>G at p<0.05 into seven anatomical sites. For example, cardiovascular disorder included non-ischaemic cardiomyopathy all cause heart failure (Table S2). Similar to diabetes, the individuals with 3-4.9% m.3243A>G levels did not have a statistically significant increased risk of having any one disorder or any two disorders (fisher exact test p>0.1). However the association became significant with increasing heteroplasmy and, compared to non-carriers, individuals with ≥10% m.3243A>G heteroplasmy had a higher chance of having any one disorder (OR 2.75, 95% CI [0.71-9.52], p=0.076), increasing to 24.8 (95% CI [8.1-75.7], p<5.1x10^-8^) for any two disorders and 53.50 (95% CI [11.78-199.93], p<3.5x10^-6^) for any three or more disorders (p heterogeneity 0.01) (Figure 2B). Although the risk of multiple features was high compared to non-carriers, it proportionally affected only a small number of total m.3243A>G carriers (n=83); 23% (n=19) with one disorder, 18% (n=15) with two disorders and 7% (n=6) with three or more, which increased with higher heteroplasmy (Figure 2C). This indicates that mutation load is an important factor in the expressivity of these multi-system traits in the unselected population.

### Age-related penetrance of diabetes is lower in unselected populations with m.3243A>G compared to clinically selected diabetes cohorts

Penetrance estimates based on clinically selected probands are often overinflated for nuclear monogenic disorders (19). However, this ascertainment effect has not been explored in detail for the m.3243A>G variant. We therefore compared the penetrance of diabetes in the UKB to 95 probands with diabetes and m.3243A>G, identified from routine diabetes clinics (Tables S19-11). The penetrance of diabetes was 96% (95% CI [90-98]) at age 50 in probands, compared to 15% (95% CI 8.5-26) for carriers in the UKB (log rank test p=3x10^-37^) (Figure S2). The penetrance remained lower at 42% (95% CI [23-69]) even when considering individuals with >10% heteroplasmy in the UKB compared to proband individuals (p=1x10^-9^). The measured m.3243A>G level was higher in probands compared to UKBB (median 7.8 vs 24.6, Figure S1). Therefore, to assess whether the difference in penetrance is explained by difference in m.3243A>G level, we conducted cox proportional hazard model after adjustment for measured m.3243A>G heteroplasmy, age at recruitment, sex, and body mass index (BMI). We found that risk of diabetes in the UKB for individuals with >10% heteroplasmy still remained lower compared to probands (adjusted HR 0.55, 95% CI [0.43-0.7], p=9x10^-7^) (Table S11). We observed consistent results using calculated age-adjusted heteroplasmy based on published equation (35) (Table S11). These data suggest that there are additional factors modify the penetrance of diabetes in addition to m.3243A>G levels.

### Type 2 diabetes genetic risk score (T2DGRS) alters penetrance of diabetes with m.3243A>G

We hypothesized that nuclear polygenic risk might modify m.3243A>G-related diabetes as seen in nuclear monogenic disorders (36). We found that the risk of any diabetes at 50 years increased from 5% (95% CI [1-29]) to 14% (95% CI [4-46]) and 29% (95% CI [14-53]) for m.3243A>G carriers for people with low, medium, and high tertiles of T2DGRS, respectively (Figure 3). This effect was in the same direction after adjusting for m.3243A>G heteroplasmy, age, sex, and mitochondrial copy number (mtCN) with the risk of diabetes in carriers increasing by 1.62-fold (95% CI [0.97-2.7], p=0.06) per 1 SD increase in T2DGRS in m.3243A>G carriers. The impact of T2DGRS on diabetes penetrance was similar to that of non-carriers (interaction p=0.9). Importantly, this borderline association was maintained with a partitioned T2DGRS representing beta cell dysfunction (HR 1.1, 95% CI [1-1.2], p=0.04) but not with other non-beta cell partitioned GRS (data not shown). This supports the current understanding that beta cell dysfunction is a primary cause of diabetes in m.3243A>G individuals (37).

**Figure 3.**
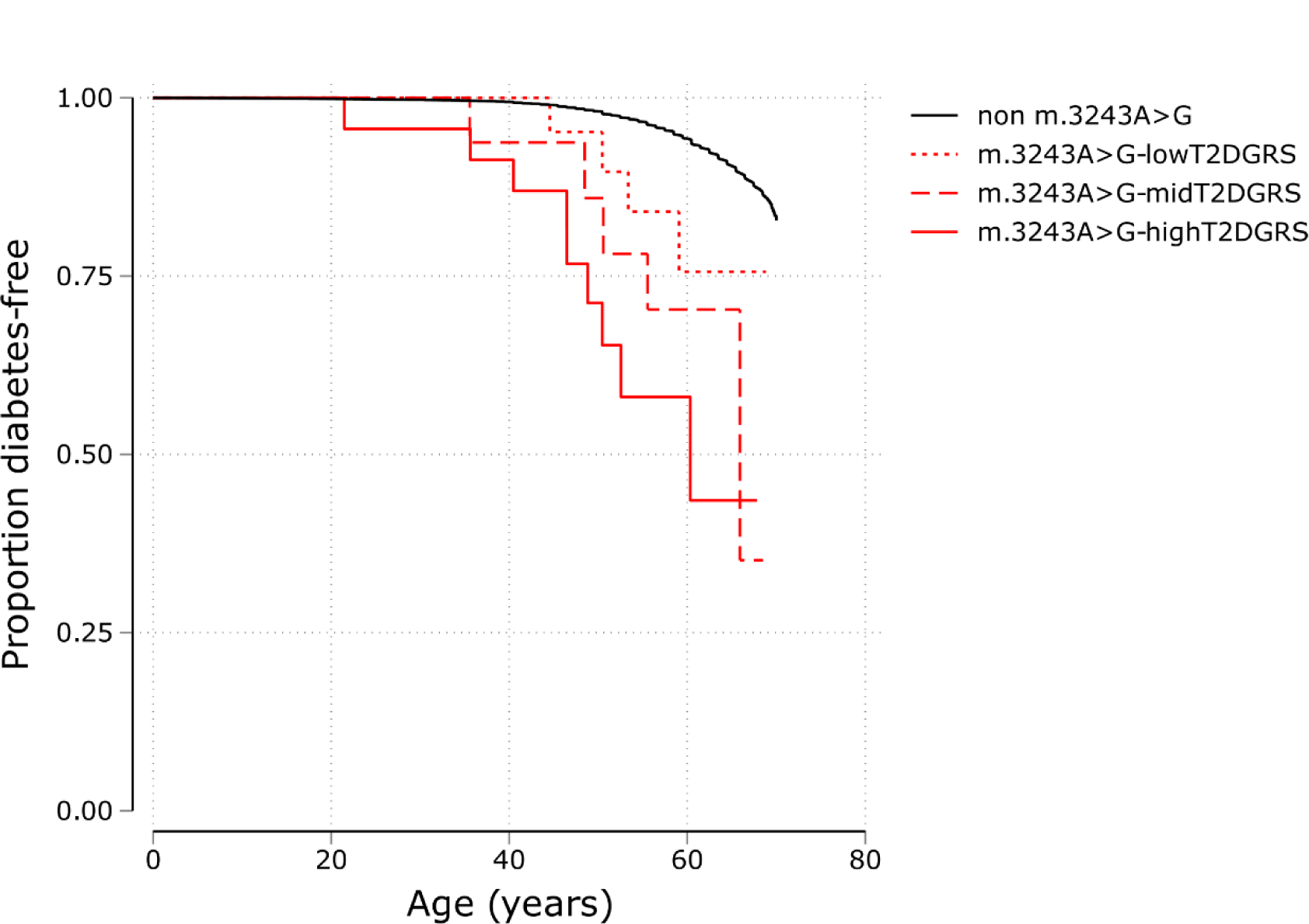
Penetrance of diabetes for individuals with pathogenic *m.3243A>G* variants in an unselected cohort. Kaplan-Meier survival curves of diabetes for m.3243A>G carriers split into tertiles (low, mid, and high) of type 2 diabetes genetic risk score (T2DGRS) and non-carriers (n=178,340) in the UK Biobank. The log rank test p value for low vs mid and low vs high groups was 0.39 and 0.03, respectively.

### Most previously reported pathogenic variants are rare in a population cohort

We next assessed the frequency and association of known pathogenic variants on our 15 mitochondrial-related traits. Of 73 well-characterised known pathogenic variants, 13 were not present in UKB (17.81%), 35 were very rare with a frequency of <1:35973 (n≤5 carriers), 17 with a frequency between 1:8994-29977 (n=6-20 carriers), five with a frequency between 1:1799-8565 (n=21-100 carriers; m.1494C>T, m.3243A>G, m.3460G>A, m.8344A>G, m.8969G>A), and three with a frequency >1:1400 (m.11778G>A, n=123 carriers; m.14484T>C, n=190 carriers; m.1555A>G, n=514 carriers) (Figure 4, Table S4). Variants causing deafness were most common at 1:263 people (n=684) followed by Leber hereditary optic neuropathy (LHON) disease-causing variants at 1:439 (n=410) although even the move common variants showed very few affected participants, except for m.3243A>G (Table 2).

**Figure 4.**
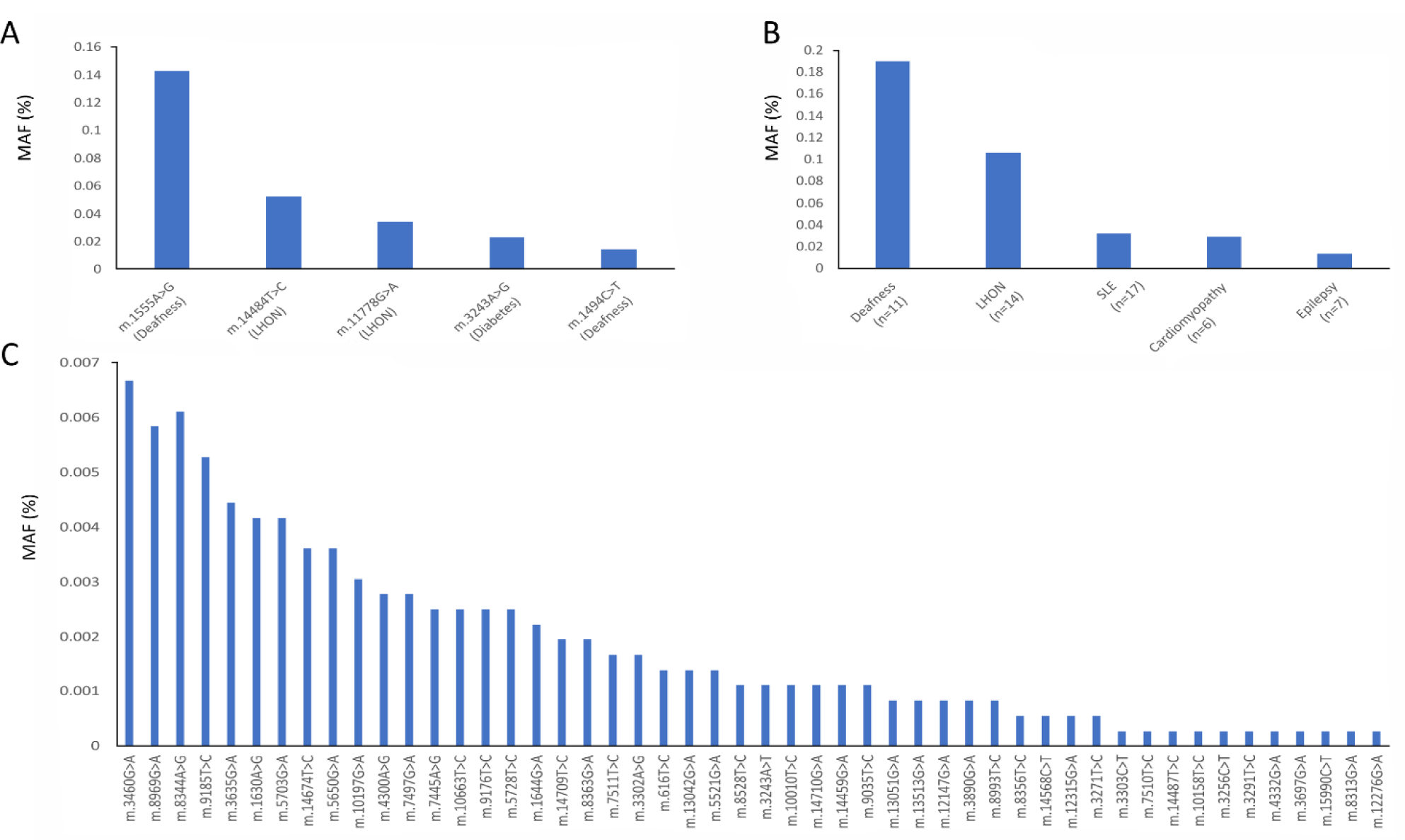
Minor allele frequency (%) of pathogenic mitochondrial variants in the UKB. **A** The five most common variants **B** Variants (n) grouped by disease **C** The remaining 50 variants present in the UK Biobank. LHON=Leber hereditary optic neuropathy; MIDD=Maternally inherited diabetes and deafness (note: mitochondrial encephalomyopathy, lactic acidosis and stroke-like episodes (MELAS) and other phenotypes are also associated with this variant); SLE=Stroke-like episodes.

**Table 2:** Penetrance of relevant phenotype in carriers of 6 most common pathogenic variants in UKB. MIDD=Maternally inherited diabetes and deafness. LHON= Leber hereditary optic neuropathy.

| Pathogenic Variant (disease) | Phenotype assessed in UKB | Phenotype frequency in carriers (%) | Phenotype frequency in non-carriers (%) |
| --- | --- | --- | --- |
| m.3243A>G (MIDD) | Diabetes | 24/83 (28.92) | 16362/179867 (0.09) |
| m.1555A>G (deafness) | Hearing aid use | 19/514 (3.7) | 6193/179438 (3.45) |
| m.14484T>C (LHON) | Bilateral visual loss | 0/190 | 124/179762 (0.07) |
| m.11778G>A (LHON) | Bilateral visual loss | 0/123 | 124/179829 (0.07) |
| m.1494C>T (Deafness) | Hearing aid use | 0/53 | 6212/179899 (3.45) |
| m.3460G>A (LHON) | Bilateral visual loss | 0/24 | 124/179928 (0.07) |
| m.8344A>G (Epilepsy) | Epilepsy | 0/22 | 2635/179929 (1.46) |

Most of the variants were too rare to perform statistically well-powered single variant analysis. However, for the seven variants (except m.3243A>G) where we had >20 individuals in the EUR-like ancestry cohort, we did not observe association with the mitochondrial-related traits at a Bonferroni-corrected threshold (p<0.0006) (Table S12). We also did not observe association with the respective traits when variants were aggregated by disease-associated phenotypes. The penetrance of relevant traits for these variants were very low and ranged from 0-3.7%.

## Discussion

Our study demonstrates that large-scale WGS data offer an exciting opportunity to study the role of mitochondria in human health. This approach can identify important mitochondrial variants and provide novel insights which may lead to the possibility of reporting mitochondrial variants discovered incidentally.

Population studies allow a unique opportunity to assess the frequency of pathogenic variants which have been previously identified in cohorts referred because of presenting diseases. The m.3243A>G variant is the most common cause of adult mitochondrial disease and due to its heteroplasmic nature it is absent from whole genome data derived from genotyping arrays. Here we used WGS data to accurately genotype m.3243A>G carriers in a population cohort of ∼180,000 participants and identified 83 cases with heteroplasmy ≥3% in blood (1 in 2167). This frequency may be an underestimate as it is well known that blood heteroplasmy reduces with age (35) and the mean age of our cohort was 56.89 years (n=179,862; SD=8.1, range 38.83-72.92). Previous attempts at identifying population frequency have been limited to small, and/or selected cohorts, with estimates ranging from 0.017-1.69% (38, 39, 40, 41, 42, 43).

We were surprised to find only one variant (m.3243A>G) associated with diabetes despite having large numbers of people with diabetes in our cohort. We observe that penetrance in the unselected population increased with m.3243A>G level, reaching 42% at age 50 for people with measured heteroplasmy of ≥10% (age adjusted heteroplasmy ∼45%). The risk ratio of diabetes at ≥10% heteroplasmy (age-adjusted heteroplasmy ∼45%) is comparable to that of pathogenic variants in well-known monogenic diabetes genes in the same cohort, as we previously published (19).

Although heteroplasmy was a major factor affecting penetrance, we also identify that polygenic risk of type 2 diabetes can also modify the penetrance. These results will need to be replicated in larger cohorts but demonstrate an exciting interplay between nuclear and mitochondrial genome variants responsible for the onset of diabetes in humans. The observation of lower penetrance in an unselected population, compared to a clinically ascertained cohort, has also been recently reported in nuclear monogenic diabetes (19) and in mitochondrial disorders such as LHON (44).

Despite the large sample size and examining 15 mitochondrial-related traits the only trait where we have shown multiple mitochondrial variants association is AST. We identified 12 mtDNA variants associated with AST levels, nine of which were previously reported in the same cohort when these variants were called from genotyping array data (13). The novel lead variant m.13488C>T also showed an association with ALT, suggesting that these variants are likely to increase AST level by their effect on the liver, rather than on the heart or skeletal muscle, which can also be sources of AST. The effect size on AST was small, and the lack of association with NAFLD suggests that the high AST level was not due to hepatocellular damage/leakage but may reflect a rise in mitochondrial AST isoform rather than cytoplasmic AST (45, 46). Mitochondrial dysfunction has been reported to play a significant role in the pathogenesis of non-alcoholic fatty liver disease (NAFLD; for review, see (47)). However, we did not identify any genome-wide significant association with NAFLD. Despite a sample size that was substantially larger, we were unable to replicate the previous borderline (p=0.06) association of m.16318A>C (p=0.38) (48). The original finding may be a false-positive due to the small sample size (n∼300), but the disparity could also be explained by less well-defined phenotype capture in UKB and a reduced effect size in a non-clinically ascertained cohort.

Ours is the largest study of mitochondrial variants based on whole genome sequence data. This provides unique opportunity to assess the population frequency of pathogenic variants which help to better understand the burden of mitochondrial disease in population. We found that most variants were rare in population in line with them causing rare mitochondrial disorders except for some variants causing deafness (m.1555A>G, MAF=0.0014; m.1494C>T, MAF=0.00014) and LHON (m.14484T>C, MAF=0.0005; m.11778G>A, MAF=0.0003). Both m.1555A>G and m.1494C>T cause deafness only after exposure to aminoglycoside antibiotics, which may explain why we failed to detect an association with deafness with either in isolation or when combined (49). We also did not observe an association with bilateral vision loss for the LHON variants (n=10) identified. This may be due to a combination of the lower sample size of bilateral vision loss in our cohort (n=140) and the well-known low penetrance of the LHON variants in the unselected population (50). Although we used mitochondrial-related traits, our phenotypes were more general and may have overshadowed the specific phenotype of mitochondrial-related diseases (such as stroke-like episodes vs. all strokes (51)). This, along with the low number of pathogenic variants and despite our large cohort, may explain the lack of association of known pathogenic variants in our study.

Our study has some limitations. Although our study was one of the largest to assess mitochondrial-wide association, we were still limited in power for non-European-like ancestry populations. Similarly, it is known that the UK Biobank has a healthy volunteer selection bias which is not fully representative of the UK population (52) and may have therefore limited the inferences we can make about multisystem disease. Our stringent sample and variant filter criteria mean that it is possible that we may have excluded true causal or associated variants with these 15 mitochondrial-related traits. However, these stringent criteria allowed us to investigate the association of low level heteroplasmy. This was particularly important for m.3243A>G, which is commonly considered to be present in blood with a heteroplasmy ≥3% (35, 53, 54). Some of the phenotypes we assessed had a lower sample size (e.g. bilateral vision loss), which will have limited our ability to robustly assess any associations and would require larger sample sizes to better perform association discovery. We also limited our analysis to single nucleotide variants and small insertions/deletions and did not assess larger deletions, which have been implicated in mitochondrial disease. Additionally, we only used heteroplasmy identified from whole blood samples, which may not reflect heteroplasmy in other tissues or organs that are more relevant to specific mitochondrial diseases.

Our study has important implications for the incidental identification of pathogenic mitochondrial variants. Diagnostic molecular genetic laboratories are moving towards using whole-genome sequencing as a first-line genetic test. This, along with a rapid rise in direct-to-consumer testing and use of WGS for research, provides an exciting opportunity to obtain information on pathogenic mitochondrial variants, well before any disease onset. We show that, when detected incidentally from blood, measured m.3243A>G heteroplasmy ≥10% (age-adjusted heteroplasmy ∼45%) significantly increases the risk of diabetes, heart failure, deafness, and that individuals with this level or greater are more likely to experience multiple system disorders. Our findings, combined with the growing availability of prenatal testing for mitochondrial disease, indicate that reporting of m.3243A>G variant when discovered incidentally could have a significant health benefit, particularly for female individuals of reproductive age. If one decides to report this variant when found incidentally, our data suggest that this may be beneficial to individuals with age-adjusted heteroplasmy levels over 45%. However, further studies in unselected population(s) will be needed to refine this advice.

## Data and Code Availability

The data supporting the findings of this study are available within the article and its Supplementary Data files. Additional information for reproducing the results described in the article is available upon reasonable request and subject to a data use agreement. The UK Biobank dataset is available from https://biobank.ctsu.ox.ac.uk.

## Acknowledgements

This research has been conducted using the UK Biobank Resource. This work was carried out under UK Biobank project number 9072. The current work is funded by Diabetes UK (19/0005994 and 21/0006335), MRC (MR/T00200X/1) and Wellcome Trust’s Institutional Strategic Support Fund awarded to University of Exeter. KAP is funded by Wellcome Trust (219606/Z/19/Z), ATH is supported by Wellcome Trust Senior Investigator award (WT098395/Z/12/Z) and SJP is funded by the Wellcome Career Re-entry Fellowship (204709/Z/16/Z) and the Wellcome Centre for Mitochondrial Research (203105/Z/16/Z). The work is supported by the National Institute for Health Research (NIHR) Exeter Biomedical Research Centre, Exeter, UK. The Wellcome Trust, MRC and NIHR had no role in the design and conduct of the study; collection, management, analysis, and interpretation of the data; preparation, review, or approval of the manuscript; and decision to submit the manuscript for publication. The views expressed are those of the author(s) and not necessarily those of the Wellcome Trust, Department of Health, NHS or NIHR. For the purpose of open access, the author has applied a CC BY public copyright licence to any Author Accepted Manuscript version arising from this submission.

## References

1. Gorman GS, Chinnery PF, DiMauro S, Hirano M, Koga Y, McFarland R, et al. Mitochondrial diseases. Nat Rev Dis Primers. 2016;2:16080.

2. Thompson K, Collier JJ, Glasgow RIC, Robertson FM, Pyle A, Blakely EL, et al. Recent advances in understanding the molecular genetic basis of mitochondrial disease. Journal of Inherited Metabolic Disease. 2020;43(1):36–50.

3. Li D, Liang C, Zhang T, Marley JL, Zou W, Lian M, et al. Pathogenic mitochondrial DNA 3243A>G mutation: From genetics to phenotype. Front Genet. 2022;13:951185.

4. Bernardino Gomes TM, Ng YS, Pickett SJ, Turnbull DM, Vincent AE. Mitochondrial DNA disorders: from pathogenic variants to preventing transmission. Hum Mol Genet. 2021;30(R2):R245–r53.

5. Brandon MC, Lott MT, Nguyen KC, Spolim S, Navathe SB, Baldi P, et al. MITOMAP: a human mitochondrial genome database--2004 update. Nucleic Acids Res. 2005;33(Database issue):D611-3.

6. Reardon W, Ross RJ, Sweeney MG, Luxon LM, Pembrey ME, Harding AE, et al. Diabetes mellitus associated with a pathogenic point mutation in mitochondrial DNA. Lancet. 1992;340(8832):1376-9.

7. Kraja AT, Liu C, Fetterman JL, Graff M, Have CT, Gu C, et al. Associations of Mitochondrial and Nuclear Mitochondrial Variants and Genes with Seven Metabolic Traits. Am J Hum Genet. 2019;104(1):112–38.

8. Calabrese C, Pyle A, Griffin H, Coxhead J, Hussain R, Braund PS, et al. Heteroplasmic mitochondrial DNA variants in cardiovascular diseases. PLOS Genetics. 2022;18(4):e1010068.

9. Hudson G, Nalls M, Evans JR, Breen DP, Winder-Rhodes S, Morrison KE, et al. Two-stage association study and meta-analysis of mitochondrial DNA variants in Parkinson disease. Neurology. 2013;80(22):2042–8.

10. Xia C, Pickett SJ, Liewald DCM, Weiss A, Hudson G, Hill WD. The contributions of mitochondrial and nuclear mitochondrial genetic variation to neuroticism. Nat Commun. 2023;14(1):3146.

11. Hudson G, Gomez-Duran A, Wilson IJ, Chinnery PF. Recent mitochondrial DNA mutations increase the risk of developing common late-onset human diseases. PLoS Genet. 2014;10(5):e1004369.

12. Ng YS, Lim AZ, Panagiotou G, Turnbull DM, Walker M. Endocrine Manifestations and New Developments in Mitochondrial Disease. Endocr Rev. 2022;43(3):583–609.

13. Yonova-Doing E, Calabrese C, Gomez-Duran A, Schon K, Wei W, Karthikeyan S, et al. An atlas of mitochondrial DNA genotype–phenotype associations in the UK Biobank. Nature Genetics. 2021;53(7):982–93.

14. Børte S, Zwart JA, Skogholt AH, Gabrielsen ME, Thomas LF, Fritsche LG, et al. Mitochondrial genome-wide association study of migraine - the HUNT Study. Cephalalgia. 2020;40(6):625–34.

15. Aboulmaouahib B, Kastenmüller G, Suhre K, Zöllner S, Weissensteiner H, Prehn C, et al. First mitochondrial genome-wide association study with metabolomics. Hum Mol Genet. 2022;31(19):3367–76.

16. Sookoian S, Flichman D, Scian R, Rohr C, Dopazo H, Gianotti TF, et al. Mitochondrial genome architecture in non-alcoholic fatty liver disease. J Pathol. 2016;240(4):437–49.

17. Han X, Souzeau E, Ong JS, An J, Siggs OM, Burdon KP, et al. Myocilin Gene Gln368Ter Variant Penetrance and Association With Glaucoma in Population-Based and Registry-Based Studies. JAMA Ophthalmol. 2019;137(1):28–35.

18. Patel AP, Dron JS, Wang M, Pirruccello JP, Ng K, Natarajan P, et al. Association of Pathogenic DNA Variants Predisposing to Cardiomyopathy With Cardiovascular Disease Outcomes and All-Cause Mortality. JAMA Cardiol. 2022;7(7):723–32.

19. Mirshahi UL, Colclough K, Wright CF, Wood AR, Beaumont RN, Tyrrell J, et al. Reduced penetrance of MODY-associated HNF1A/HNF4A variants but not GCK variants in clinically unselected cohorts. Am J Hum Genet. 2022;109(11):2018–28.

20. Laricchia KM, Lake NJ, Watts NA, Shand M, Haessly A, Gauthier L, et al. Mitochondrial DNA variation across 56,434 individuals in gnomAD. Genome Res. 2022;32(3):569–82.

21. Halldorsson BV, Eggertsson HP, Moore KHS, Hauswedell H, Eiriksson O, Ulfarsson MO, et al. The sequences of 150,119 genomes in the UK Biobank. Nature. 2022;607(7920):732-40.

22. Venner E, Muzny D, Smith JD, Walker K, Neben CL, Lockwood CM, et al. Whole-genome sequencing as an investigational device for return of hereditary disease risk and pharmacogenomic results as part of the All of Us Research Program. Genome Medicine. 2022;14(1):34.

23. González MDM, Ramos A, Aluja MP, Santos C. Sensitivity of mitochondrial DNA heteroplasmy detection using Next Generation Sequencing. Mitochondrion. 2020;50:88–93.

24. Bycroft C, Freeman C, Petkova D, Band G, Elliott LT, Sharp K, et al. The UK Biobank resource with deep phenotyping and genomic data.

25. Ng YS, Bindoff LA, Gorman GS, Klopstock T, Kornblum C, Mancuso M, et al. Mitochondrial disease in adults: recent advances and future promise. Lancet Neurol. 2021;20(7):573–84.

26. Battle SL, Puiu D, Group TOmW, Verlouw J, Broer L, Boerwinkle E, et al. A bioinformatics pipeline for estimating mitochondrial DNA copy number and heteroplasmy levels from whole genome sequencing data. NAR Genomics and Bioinformatics. 2022;4(2):lqac034.

27. Benjamin D, Sato T, Cibulskis K, Getz G, Stewart C, Lichtenstein L. Calling Somatic SNVs and Indels with Mutect2. bioRxiv. 2019:861054.

28. Mn W, L J, Jw H, Ks R, J T, At H, et al. Use of SNP chips to detect rare pathogenic variants: retrospective, population based diagnostic evaluation. BMJ. 2021;372:n214.

29. Mahajan A, Go MJ, Zhang W, Below JE, Gaulton KJ, Ferreira T, et al. Genome-wide trans-ancestry meta-analysis provides insight into the genetic architecture of type 2 diabetes susceptibility. Nat Genet. 2014;46(3):234–44.

30. Morris AP, Voight BF, Teslovich TM, Ferreira T, Segrè AV, Steinthorsdottir V, et al. Large-scale association analysis provides insights into the genetic architecture and pathophysiology of type 2 diabetes. Nat Genet. 2012;44(9):981–90.

31. Udler MS, Kim J, von Grotthuss M, Bonàs-Guarch S, Cole JB, Chiou J, et al. Type 2 diabetes genetic loci informed by multi-trait associations point to disease mechanisms and subtypes: A soft clustering analysis. PLoS Med. 2018;15(9):e1002654.

32. Singh R, Ellard S, Hattersley A, Harries LW. Rapid and sensitive real-time polymerase chain reaction method for detection and quantification of 3243A>G mitochondrial point mutation. J Mol Diagn. 2006;8(2):225–30.

33. Mbatchou J, Barnard L, Backman J, Marcketta A, Kosmicki JA, Ziyatdinov A, et al. Computationally efficient whole-genome regression for quantitative and binary traits. Nature Genetics. 2021;53(7):1097–103.

34. McLaren W, Gil L, Hunt SE, Riat HS, Ritchie GRS, Thormann A, et al. The Ensembl Variant Effect Predictor. Genome Biology. 2016;17(1):122.

35. Grady JP, Pickett SJ, Ng YS, Alston CL, Blakely EL, Hardy SA, et al. mtDNA heteroplasmy level and copy number indicate disease burden in m.3243A>G mitochondrial disease. EMBO Molecular Medicine. 2018;10(6):e8262.

36. Khera AV, Chaffin M, Aragam KG, Haas ME, Roselli C, Choi SH, et al. Genome-wide polygenic scores for common diseases identify individuals with risk equivalent to monogenic mutations. Nat Genet. 2018;50(9):1219–24.

37. Lindroos MM, Majamaa K, Tura A, Mari A, Kalliokoski KK, Taittonen MT, et al. m.3243A>G mutation in mitochondrial DNA leads to decreased insulin sensitivity in skeletal muscle and to progressive beta-cell dysfunction. Diabetes. 2009;58(3):543-9.

38. Majamaa K, Moilanen JS, Uimonen S, Remes AM, Salmela PI, Kärppä M, et al. Epidemiology of A3243G, the mutation for mitochondrial encephalomyopathy, lactic acidosis, and strokelike episodes: prevalence of the mutation in an adult population. Am J Hum Genet. 1998;63(2):447-54.

39. Elliott HR, Samuels DC, Eden JA, Relton CL, Chinnery PF. Pathogenic mitochondrial DNA mutations are common in the general population. Am J Hum Genet. 2008;83(2):254–60.

40. Manwaring N, Jones MM, Wang JJ, Rochtchina E, Howard C, Mitchell P, et al. Population prevalence of the MELAS A3243G mutation. Mitochondrion. 2007;7(3):230–3.

41. Odawara M, Sasaki K, Yamashita K. Prevalence and clinical characterization of Japanese diabetes mellitus with an A-to-G mutation at nucleotide 3243 of the mitochondrial tRNA(Leu(UUR)) gene. J Clin Endocrinol Metab. 1995;80(4):1290–4.

42. Bouhaha R, Abid Kamoun H, Elgaaied A, Ennafaa H. A3243G mitochondrial DNA mutation in Tunisian diabetic population. Tunis Med. 2010;88(9):642-5.

43. Wang S, Wu S, Zheng T, Yang Z, Ma X, Jia W, et al. Mitochondrial DNA mutations in diabetes mellitus patients in Chinese Han population. Gene. 2013;531(2):472–5.

44. Mackey DA, Ong JS, MacGregor S, Whiteman DC, Craig JE, Lopez Sanchez MIG, et al. Is the disease risk and penetrance in Leber hereditary optic neuropathy actually low? Am J Hum Genet. 2023;110(1):170–6.

45. Jiang X, Chang H, Zhou Y. Expression, purification and preliminary crystallographic studies of human glutamate oxaloacetate transaminase 1 (GOT1). Protein Expr Purif. 2015;113:102–6.

46. Glinghammar B, Rafter I, Lindström AK, Hedberg JJ, Andersson HB, Lindblom P, et al. Detection of the mitochondrial and catalytically active alanine aminotransferase in human tissues and plasma. Int J Mol Med. 2009;23(5):621–31.

47. Dabravolski SA, Bezsonov EE, Baig MS, Popkova TV, Nedosugova LV, Starodubova AV, et al. Mitochondrial Mutations and Genetic Factors Determining NAFLD Risk. Int J Mol Sci. 2021;22(9).

48. Hasturk B, Yilmaz Y, Eren F. Potential clinical variants detected in mitochondrial DNA D-loop hypervariable region I of patients with non-alcoholic steatohepatitis. Hormones (Athens). 2019;18(4):463–75.

49. Ealy M, Lynch KA, Meyer NC, Smith RJH. The prevalence of mitochondrial mutations associated with aminoglycoside-induced sensorineural hearing loss in an NICU population. The Laryngoscope. 2011;121(6):1184–6.

50. Lopez Sanchez MIG, Kearns LS, Staffieri SE, Clarke L, McGuinness MB, Meteoukki W, et al. Establishing risk of vision loss in Leber hereditary optic neuropathy. Am J Hum Genet. 2021;108(11):2159–70.

51. Ng YS, Bindoff LA, Gorman GS, Horvath R, Klopstock T, Mancuso M, et al. Consensus-based statements for the management of mitochondrial stroke-like episodes. Wellcome Open Res. 2019;4:201.

52. Fry A, Littlejohns TJ, Sudlow C, Doherty N, Adamska L, Sprosen T, et al. Comparison of Sociodemographic and Health-Related Characteristics of UK Biobank Participants With Those of the General Population. Am J Epidemiol. 2017;186(9):1026–34.

53. Mavraki E, Labrum R, Sergeant K, Alston CL, Woodward C, Smith C, et al. Genetic testing for mitochondrial disease: the United Kingdom best practice guidelines. Eur J Hum Genet. 2023;31(2):148–63.

54. de Laat P, Janssen MC, Alston CL, Taylor RW, Rodenburg RJ, Smeitink JA. Three families with ’de novo’ m.3243A > G mutation. BBA Clin. 2016;6:19-24.

