## Supplementary Data files for "Large-scale blood mitochondrial genome-wide study provides novel insights into mitochondrial disease-related traits"

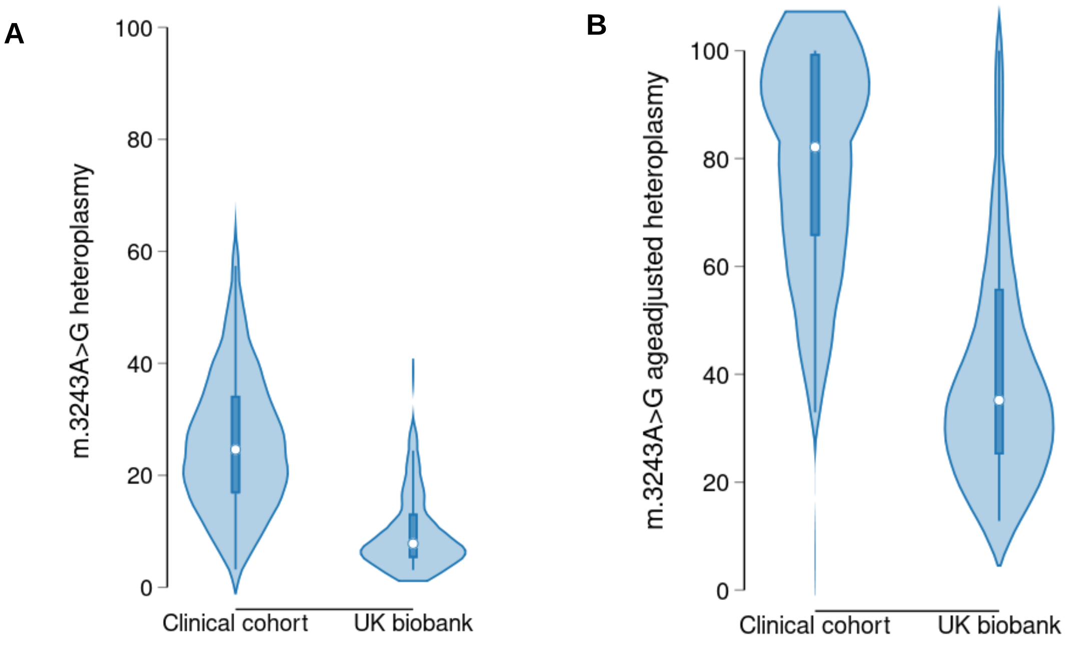


**Figure S1: Measured and age-adjusted *m.3243A>G* heteroplasmy in clinically selected and unselected cohorts.** Violin plots showing **A** measured heteroplasmy and **B** predicted age-adjusted heteroplasmy (for formula see: [1]). Clinically ascertained cases were referred with diabetes and found to have the m.3243A>G variant (n=95). Clinically unselected cases were obtained from the UK Biobank population cohort (n=83).

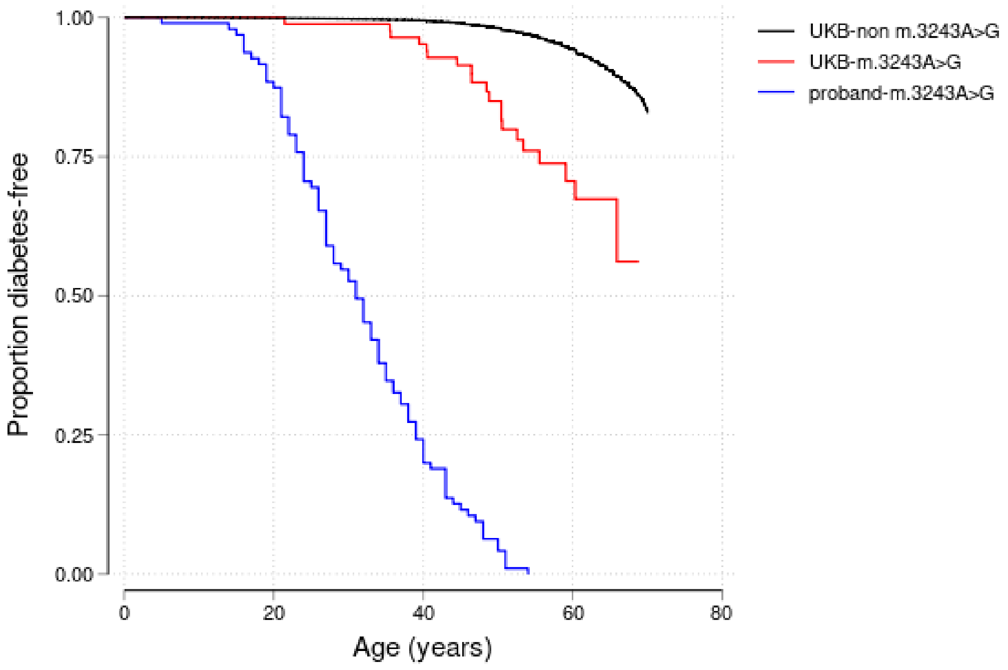


**Figure S2:**

**Combined penetrance of diabetes for individuals with pathogenic *m.3243A>G* variants in clinically selected and unselected cohorts.** Kaplan-Meier survival curves of diabetes for *m.3243A>G* clinically identified probands (N=95) and individuals from UK Biobank population cohort (n=83) and individuals from UK Biobank population cohort without *m.3243A>G* variant (n=179,340). The log rank test p value for UK biobank carriers against probands was $3x10^{-37}$ and against UK Biobank noncarriers was $2x10^{-19}$.

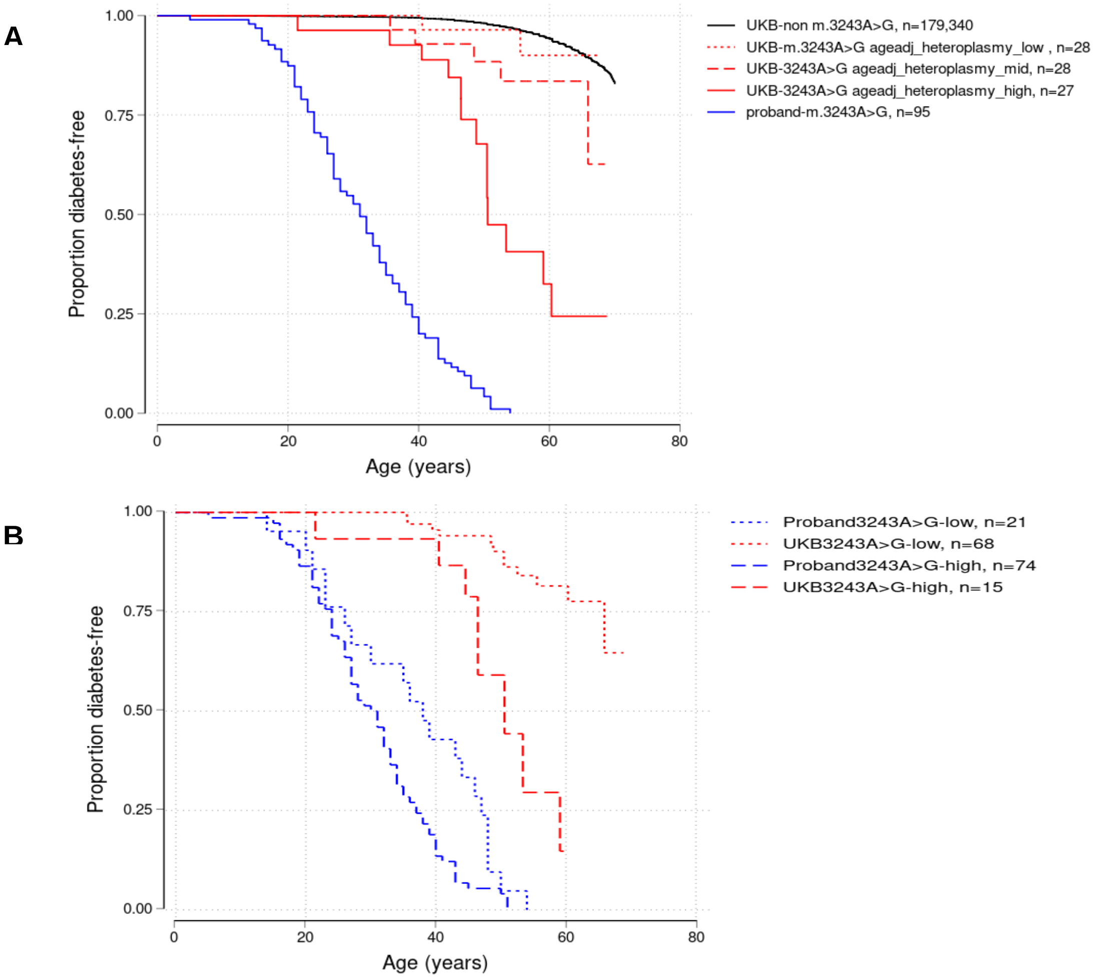


**Figure S3: Penetrance of diabetes for individuals with pathogenic *m.3243A>G* variants in clinically selected and unselected cohorts using age-adjusted heteroplasmy categories**. **A** Kaplan-Meier survival curves of diabetes for m.3243A>G probands (N=95) and carriers from the UK Biobank population cohort split into tertiles (low, mid, and high) of age adjusted heteroplasmy level (for formula see: [1]). The log rank test p value of highest tertile against proband was $6x10^{-12}$. The penetrance at age 50 for higher tertile group was 33% (95% CI 16-57) **B** Kaplan-Meier survival curves split by median of the age adjusted heteroplasmy of the combined probands and m.3243A>G carriers from the UK Biobank (<64% vs >64%). The log rank test p value for low heteroplasmy groups from UK biobank and probands was $2x10^{-22}$and was $1x10^{-07}$ for high heteroplasmy groups.

1. Grady, J.P., et al., *mtDNA heteroplasmy level and copy number indicate disease burden in m.3243A>G mitochondrial disease.* EMBO Molecular Medicine, 2018. **10**(6): p. e8262.
