## Supplementary Data files for "Large-scale blood mitochondrial genome-wide study provides novel insights into mitochondrial disease-related traits"

| Characteristic | m.3243 A>G from unselected UK Biobank cohort (n=83) | m.3243 A>G probands – clinically referred with suspected mitochondrial diabetes (n=95) |
|---|---|---|
| Age (years) | 55.3 (47-62) | 39 (31-48) |
| Sex | 41 (49%) | 32 (34%) |
| BMI (kg/m$^2$) | 24.3 (21.7-26.9) | 22 (20-24.5) |
| Diabetes (baseline and follow up) | 24 (29%) | 100% |
| Diabetes at baseline | 20 (24%) | 100% |
| Age at diabetes (years) | 48.5 (40-52.5) | 31 (24-39) |
| Deafness* | 20 (24%) | 50 (53%) |
| Mother with diabetes | 10 (13%) | 68 (78%) |
| Insulin treatment | 7/20 (35%) | 65 (73%) |
| m.3243A>G heteroplasmy | | |
| Overall (%) | 7.8 (5.3-13.1) | 24.6 (16.8-34.2) |
| 3-4.9 (%) | 19 (23%) | 1 (1%) |
| 5-9.9 (%) | 39 (47%) | 5 (5%) |
| ≥10 (%) | 25 (30%) | 89 (94%) |
| Overall age adjusted (%) | 35 (25-59) | 82(66-100) |

**Table S2 - Characteristics of m.3243A>G carriers from the UK Biobank and probands from a clinically referred cohort.** m 3243A>G heteroplasmy reduces with age. We therefore, used published regression equation to calculate the age-adjusted heteroplasmy for our cohort [1]. Values are median (IQR) for continuous variables or number of individuals (%) for categorical variables. The number of individuals with available data is also indicated where appropriate. *Clinician reported deafness in the proband (no data on hearing aid available), the corresponding number in the UK biobank refers to people using hearing aid as a marker for severe hearing loss. BMI=body mass index.

| Clinical Phenotype | Definition |
|---|---|
| **All-cause heart failure** | Self-reported history of heart failure or cardiomyopathy during verbal interview with trained nurse; or ICD10 code from hospitalization or death records for hypertensive heart disease, cardiomyopathy or heart failure (I11.0, I13.0, I13.2, I25.5, I42.0, I42.5, I42.8, I42.9, I50.0, I50.1, I50.9); or hospitalization due to ICD-9 code for heart failure or other primary cardiomyopathies (4254, 4280, 4281, 4289); excluding individuals with history of hypertrophic cardiomyopathy during verbal interview with trained nurse, or hospitalization for or death due to ICD-10 code for hypertrophic cardiomyopathy (I42.1, I42.2) |
| **Nonischemic cardiomyopathy** | ICD10 code from Hospitalization for or death records for dilated cardiomyopathy or left ventricular failure (I42.0, I50.1); **or** Hospitalization due to ICD-9 code for left heart failure (4281); or history of hypertrophic cardiomyopathy during verbal interview with trained nurse, or hospitalization for or death due to ICD-10 code for hypertrophic cardiomyopathy (I42.1, I42.2) excluding individuals with history of coronary artery, |
| **Nonischemic arrythmia** | Self-reported history of any arrythmia, or cardioversion during verbal interview with trained nurse; **or** hospitalization for or death due to ICD-10 code for atrial fibrillation or atrial flutter (I44 I45 I47 I48 I49); **or** hospitalization due to ICD-9 code for atrial fibrillation or atrial flutter (426x, |

| | |
|---|---|
| | 427x) or operational code for ablation (X50 K571 K572 K575 K576 K601 K605 K611). Excluding individuals with history of coronary artery |
| **Hearing aid use** | Self-reported history during verbal interview with trained nurse; Hospitalization for or death due to ICD-10 code for hearing aid use (Z461 Z453) |
| **Sensory neural deafness** | ICD-10 code for sensory neural hearing loss (H903 H904 H905 H919) from Hospitalization or death records excluding the cases with nonsensory neural deafness from ICD10 code from hospitalazisation and death (H900 H901 H902 H906 H907 H910 H911 H912 H913 H918 Q16) |
| **Bilateral vision loss** | ICD-10 code for sensory neural hearing loss (H542 H543 H549 H47.2) from Hospitalization or death records or missed OCT or visual acuity testing due to visually impaired in both eye, **excluding** people with congenital blindness using ICD10 code (Q14 Q13 Q15) and self report blindness due to glaucoma, diabetes, injury and macular degeneration |
| **Constipation** | Self-reported history of constipation or taking constipation medication during verbal interview with trained nurse; **or ICD10 code from** hospitalization or death records for hypertensive heart disease, cardiomyopathy or heart failure (K590); **or** hospitalization due to ICD-9 code for heart failure or other primary cardiomyopathies (56402 56409) |
| **Stroke** | ICD10 code from hospitalization or death records for stroke (I60, I61, I63, I64); ICD9 code from hospitalization or death records for stroke (430, 431, 434, 436); Self-reported history of stroke during verbal interview with trained nurse (1583, 1081, 1086, 1491) |
| **Epilepsy** | ICD10 code from hospitalization or death records for epilepsy (G40); ICD9 code from hospitalization or death records for epilepsy (345); Self-reported history of stroke during verbal interview with trained nurse (1264) |
| **Alanine aminotransferase (ALT)** | UK Biobank field 30620 |
| **Aspartate aminotransferase (AST)** | UK Biobank field 30650 |
| **Non-alcoholic fatty liver disease (NAFLD)** | ICD10 code from hospitalization or death records for Fatty liver disease (K76); ICD9 code from hospitalization or death records for Fatty liver disease (5715, 5716, 5718, 5719) **excluding** IC10 code from hospitalization or death records for alcoholic liver (K70); ICD9 code from hospitalization or death records for alcoholic liver (5712, 5713); Self-reported history of stroke during verbal interview with trained nurse (1604). |
| **Estimated glomerular filtration rate (eGFR; ml/min/1.73m$^2$)** | Estimated glomerular filtration rate (eGFR ml/min/1.73m$^2$) was calculated using the chronic kidney disease epidemiology collaboration (CKD-EPI) equation Creatinine-Cystatin 2012 using UK creatinine and cystatin-C from UK Biobank fields 30700 and 30720, respectively. |
| **Moderate to severe renal disease (Chronic kidney disease (CKD) plus eGFR < 45)** | ICD10 code from hospitalization of death records implicated in end-stage renal disease, reliance on dialysis, recipient of a kidney transplant plus eGFR <45 (as above): end-stage renal disease (N180), CKD stage 5 (N185), kidney transplant status (Z940), complications of kidney transplant (T861), dialysis (Z992 Z492, Z491, Z490) |
| **Diabetes** | Self report, diabetes medication, first occurrence data (ICD E10, E11, E13, E14) and HbA1c >=48 mmol/mol |
| **Multisystem phenotypes** | The seven multisystem phenotypes consisted of the above defined phenotypes but included as individuals having one, two or three or more defined as:<br><br>1. Cardiac (Nonischaemic cardiomyopathy plus all cause heart failure)<br>2. Auditory (Sensory neural deafness plus hearing aid use)<br>3. Gastrointestinal (Constipation)<br>4. Endocrine (Diabetes)<br>5. Stroke |

6. Kidneys (Moderate to severe renal disease)
7. Neurological (Epilepsy)

**Table S3** - Phenotype definitions of 15 mitochondrial disease relevant traits in the UK Biobank

| | Hazard Ratio (95% CI) | P Value | N |
|---|---|---|---|
| **All individuals** | | | |
| **Non m.3243A>G** | Base | | 179340 |
| **m.3243A>G - ALL** | 5.8 (3.8-9) | 3.5e-15 | 83 |
| **m.3243A>G - 3-4.9% heteroplasmy** | 0.98 (0.14 – 6.9) | 9.8e-01 | 19 |
| **m.3243A>G - 5-9.9% heteroplasmy** | 3.8 (1.8 – 8.0) | 3.9e-04 | 39 |
| **m.3243A>G - ≥10% heteroplasmy** | 20.6 (11.7 – 36.4) | 1.2e-25 | 25 |
| **Unrelated Europeans** | | | |
| **Non m.3243A>G** | Base | | 139016 |
| **m.3243A>G - ALL** | 6.9 (4.4-11) | 2.3e-16 | 64 |
| **m.3243A>G - 3-4.9% heteroplasmy** | 1.1 (0.15 – 7.7) | 9.4e-01 | 17 |
| **m.3243A>G - 5-9.9% heteroplasmy** | 4.9 (2.2 – 10.9) | 1.0e-04 | 26 |
| **m.3243A>G - ≥10% heteroplasmy** | 25 (13.8 – 45.2) | 2.0e-26 | 21 |

**Table S9 - Cox proportional hazard ratios for diabetes for individuals with m.3243A>G variants relative to non-carriers in the UK Biobank.** Hazard ratios (95% CI), p values, and number of all individuals used in the analysis is shown. Analysis only included prevalent diabetes cases. The model was adjusted for age at recruitment, sex, the first 10 genetic principal components, and mitochondrial copy number. Abbreviations: CI=confidence intervals.

| | Hazard Ratio (95% CI) | P Value | N |
|---|---|---|---|
| **All individuals** | | | |
| **Non m.3243A>G** | Base | | 179340 |
| **m.3243A>G - ALL** | 5.8 (3.8-9) | 3.5e-15 | 83 |
| **m.3243A>G age-adjusted heteroplasmy – low tertile (12.9-29.3%)** | 1.56 (0.4 – 6.2) | 5.3e-01 | 28 |
| **m.3243A>G age-adjusted heteroplasmy – mid tertile (29.4-45.1%)** | 3.9 (1.6 – 9.3) | 2.5e-03 | 28 |
| **m.3243A>G age-adjusted heteroplasmy – high tertile (>45.2%)** | 15.2 (8.8 – 26.1) | 1.2e-22 | 27 |
| **Unrelated Europeans** | | | |
| **Non m.3243A>G** | Base | | 139016 |
| **m.3243A>G - ALL** | 6.9 (4.4-11) | 2.3e-16 | 64 |
| **m.3243A>G age-adjusted heteroplasmy – low tertile (12.9-29.3%)** | 0.95 (0.13 – 6.7) | 9.6e-01 | 22 |
| **m.3243A>G age-adjusted heteroplasmy – mid tertile (29.4-45.1%)** | 4.4 (1.6 – 11.8) | 3.0e-03 | 19 |
| **m.3243A>G age-adjusted heteroplasmy – high tertile (>45.2%)** | 20.4 (11.8 – 35.2) | 2.1e-27 | 23 |

**Table S10 - Cox proportional hazard ratios for diabetes for individuals with m.3243A>G variants relative to non-carriers in the UK Biobank using age-adjusted heteroplasmy tertiles**. m 3243A>G heteroplasmy reduces with age. We therefore, used published regression equation to calculate the age-adjusted heteroplasmy for our cohort and split into tertiles [1] Hazard ratios (95% CI), p values, and number of all individuals used in the analysis is shown. Analysis only included prevalent diabetes cases. The model was adjusted for age at recruitment, sex, first 10 principal component, mitochondrial copy number. Abbreviations CI, confidence intervals

| | Hazard Ratio (95% CI) | P Value | N |
|---|---|---|---|
| **Unadjusted** | | | |
| **Proband m.3243A>G** | Base | | 95 |
| **UKBB m.3243A>G - ALL** | 0.05 (0.03-0.95) | 8.5e-24 | 83 |
| **UKBB m.3243A>G >10% heteroplasmy** | 0.56 (0.45 – 0.69) | 8.8e-08 | 25 |
| **UKBB m.3243A>G age-adjusted heteroplasmy – high tertile (>45.2%)** | 0.14 (0.07 – 0.26) | 2.8e-09 | 27 |
| **Adjusted** | | | |
| **Proband m.3243A>G** | Base | | 78 |
| **UKBB m.3243A>G - ALL** | 0.08 (0.04-0.15) | 1.1e-12 | 81 |
| **UKBB m.3243A>G - ALL age adjusted heteroplasmy** | 0.09 (0.04-0.17) | 8.4e-12 | 81 |
| **UKBB m.3243A>G >10% heteroplasmy** | 0.55 (0.43 – 0.7) | 9.0e-07 | 24 |
| **m.3243A>G age-adjusted heteroplasmy – high tertile (>45.2%)** | 0.17 (0.08 – 0.35) | 1.6e-06 | 26 |

**Table S11 - Cox proportional hazard ratios for diabetes for individuals with m.3243A>G from the UK biobank relative to clinically selected probands.** Hazard ratios (95% CI), p values, and number of all individuals used in the analysis is shown. Analysis only included prevalent diabetes cases. The model was adjusted for age at recruitment, sex, body mass index, heteroplasmy. Abbreviations CI, confidence intervals.

1. Grady, J.P., et al., *mtDNA heteroplasmy level and copy number indicate disease burden in m.3243A>G mitochondrial disease.* EMBO Molecular Medicine, 2018. **10**(6): p. e8262.
